# Material Hardship, Parenting, and Epigenetic Aging: A Family Stress Model of Biological Embedding in Childhood

**DOI:** 10.64898/2026.11.13.26368032

**Authors:** Georgette Metrailer, Justin Parent

## Abstract

Early-life socioeconomic adversity is linked to accelerated epigenetic aging, but how it becomes biologically embedded remains unclear. Using the Future of Families and Child Wellbeing Study (N = 1971), we tested whether material hardship was indirectly associated with children’s epigenetic aging through parenting stress and parenting behaviors. Material hardship at age 3 was associated with elevated parenting stress, which predicted greater hostility and behavioral control and lower warmth at age 5. Hostility was associated with accelerated epigenetic aging and a faster pace of aging at age 9, whereas greater warmth and behavioral control were associated with a slower pace of aging. Hardship had no direct effect on aging, but indirect pathways emerged through stress and parenting behaviors. Findings extend the Family Stress Model to molecular indicators of aging and suggest that socioeconomic adversity may become biologically embedded, in part, through interconnected family processes that confer both biological risk and protection.

---

Material hardship represents one of the most salient forms of early-life adversity (Mersky et al., 2021) and often shapes the organizing structure of children’s early environmental experiences (Conger, 1981; Gagnon et al., 2026; Rodems & Shaefer, 2020). Unlike income-based indicators of socioeconomic status, material hardship directly measures, rather than infers, a household’s lived experience of poverty by assessing a household’s ability to afford necessities such as food, stable housing, utilities, and medical care (Liu et al., 2024). Exposure to significant material hardship may be especially consequential in early childhood, as the body undergoes rapid developmental changes that are uniquely sensitive to physiological stress exposure (Gabard-Durnam & McLaughlin, 2019; Sisk & Gee, 2022). Material hardship may influence long-term health partly through dysregulation of stress-response systems such as the hypothalamic-pituitary-adrenal axis, systemic inflammation, and cardiometabolic functioning (Oh et al., 2018; Wesarg et al., 2020), and has been consistently associated with long-term physical and mental health problems (Gagnon et al., 2026; Rodems & Shaefer, 2020). Yet, much of this work has focused on direct associations between material hardship and developmental outcomes, often under examining the proximal processes through which material hardship shapes children’s daily experiences.

Parenting represents one of the most proximal and developmentally salient contexts through which children experience, interpret, and regulate stress (Darling & Steinberg, 1993; Taraban & Shaw, 2018). Thus, material hardship may become biologically embedded not only through children’s direct exposure to deprivation, but also through its effects on the caregiving environment. The Family Stress Model provides a useful framework for understanding this process, proposing that economic pressure strains caregivers’ psychological and emotional resources which increases parenting-related stress and subsequently shapes caregiving behavior and children’s developmental outcomes (Conger, 1981; Masarik & Conger, 2017). Although this model has been widely used to explain children’s behavioral and emotional adjustment (Masarik & Conger, 2017), it has yet to be extended into biological aging outcomes. This is an important omission because parenting behaviors may function as repeated stress-regulatory experiences through which children learn whether their environments are predictable, threatening, or supportive. Thus, parenting may represent a modifiable leverage point in the biological embedding of stress in early childhood.

## Epigenetic Aging

One approach to examining the biological embedding of environmental stress is through epigenetics, which refers to biochemical modifications that regulate gene expression without altering the underlying DNA sequence (Aristizabal et al., 2020; Horvath, 2013; Kusters & Horvath, 2025). Unlike static genetic variation, epigenetic patterns are dynamic and reflect both intrinsic developmental processes and interactions between biological systems and lived experience (Aristizabal et al., 2020). One of the most studied epigenetic mechanisms is DNA methylation, a process through which methyl groups attach to specific regions of DNA (CpG sites) and influence which genes are more likely to be expressed (e.g., “turned on or off”) (Horvath, 2013; Kusters & Horvath, 2025). Because DNA methylation profiles shift in systematic and highly predictable ways across the lifespan, they can be used to estimate biological age through epigenetic “clocks” (Horvath, 2013; B. S. McEwen, 2013; Raffington, 2024) which are increasingly recognized as informative biomarkers of aging and age-related biological processes (Kusters & Horvath, 2025).

Epigenetic clocks differ in how they operationalize biological aging and therefore should not be treated as interchangeable measures of a single aging process. Broadly, these clocks compare an individual’s methylation profile to normative age-related patterns (Horvath, 2013), but they operationalize biological aging in conceptually distinct ways. These differences reflect the biological processes on which they were trained, the developmental periods for which they are most appropriate, the population contexts in which they were derived and validated, and their clinical utility as biomarkers of aging-related risk (Horvath, 2026; Sehgal et al., 2026). Recent evidence demonstrates that these distinctions have meaningful consequences for biomarker performance. Across 51 longitudinal intervention studies, second-generation epigenetic biomarkers trained to capture mortality risk or pace of aging were more consistently responsive to interventions than first-generation clocks designed primarily to predict chronological age, with technically reliable measures showing particularly strong responsiveness (Sehgal et al., 2026). These findings suggest that epigenetic aging measures may capture distinct dimensions of biological change rather than interchangeable estimates of a unitary aging process.

Age-based and pace-based measures illustrate this distinction. For instance, epigenetic age acceleration (EAA) reflects the extent to which an individual’s estimates biological age differs from their chronological age, such that positive age acceleration indicates an older-than-expected methylation profile (Horvath, 2013). In contrast, pace-of-aging indices estimate the rate at which physiological systems are changing over time rather than a deviation from chronological age (Belsky et al., 2022). Thus, higher pace-of-aging estimates reflect a faster rate of biological change relative to the reference population on which the measure was calibrated (Belsky et al., 2022). Importantly, although these biomarkers are associated with morbidity, mortality, and other aging-related outcomes, no epigenetic clock has yet been established as a validated surrogate endpoint for clinical aging outcomes (Horvath, 2026). Accordingly, epigenetic aging indices are best conceptualized as complementary biomarkers of underlying biological processes rather than deterministic measures of health or aging.

## Epigenetic Aging and Material Hardship

A growing body of research suggests that adverse and socioeconomically disadvantaged early environments are associated with variation in epigenetic aging across the lifespan. Meta-analytic and longitudinal evidence indicates that greater exposure to childhood adversity is associated with EAA and faster pace-of-aging estimates, although associations vary across the type of adversity and epigenetic measure examined (Marini et al., 2020; McCrory et al., 2022; Russell et al., 2026). Early-life socioeconomic disadvantage has likewise been associated with altered epigenetic aging in children and adults (Raffington et al., 2021; Reed et al., 2026). However, much of this literature relies on broad indicators of socioeconomic adversity rather than material hardship specifically, leaving less understood about how families’ lived experiences of unmet basic needs become biologically embedded.

Emerging developmental research further suggests that variation in epigenetic aging is detectable early in the lifespan. Studies applying pediatric epigenetic clocks, such as the Pediatric-Buccal-Epigenetic (PedBE) clock, indicate that biological aging trajectories may begin to diverge in childhood and may be sensitive to variation in children’s environmental experiences (McEwen et al., 2019; Raffington et al., 2021). Yet, despite growing evidence that adversity is associated with EAA, less is known about the proximal developmental processes through which these associations emerge. This limitation is especially important because broad indicators of adversity may obscure the family-level mechanisms through which stress is repeatedly experienced, regulated, and biologically embedded.

## Parenting Under Material Hardship

Parenting under conditions of material hardship is unlikely to change uniformly across caregiving domains. Consistent with the Family Stress Model, economic pressure may increase parenting stress by straining caregivers’ emotional and cognitive resources, thereby altering the quality of everyday parent-child interactions (Masarik & Conger, 2017). Under conditions of financial instability, competing demands, and limited support, elevated parenting stress may make warm, responsive caregiving more difficult to sustain and may increase the likelihood of hostile or punitive responses during parent-child conflict (Deater-Deckard, 1998; Niu et al., 2018). At the same time, stress may not uniformly reduce behavioral control. Behavioral control refers to non-harsh efforts to structure children’s behavior through rules, monitoring, explanations, consequences, and limit-setting and is conceptually distinct from psychologically controlling or coercive parenting (Grolnick & Pomerantz, 2009; Grusec & Davidov, 2010). In disadvantaged or unpredictable environments, maintaining behavioral structure may serve a protective or adaptive function by helping caregivers establish clear expectations and promote children’s safety within otherwise constrained contexts (Brody & Flor, 1998; White et al., 2013). More broadly, parenting reflects not only parents’ responses to environmental stress but also a central socialization process through which children develop expectations about their environments and learn how to regulate their emotions and behavior within them (Grusec & Davidov, 2010).

Distinguishing warmth, behavioral control, and hostility is important because these dimensions may provide qualitatively different stress-regulatory experiences for children. Warm, responsive caregiving can promote emotional security and support children’s developing capacity for emotional and physiological co-regulation (Eisenberg et al., 2005; Grusec & Davidov, 2021). Behavioral control, when implemented without harshness or psychological coercion, may support regulation by providing clear expectations, predictable contingencies, and external scaffolding for children’s developing self-control (Grolnick & Pomerantz, 2009; Grusec & Davidov, 2021). Parental hostility, in contrast, may increase children’s exposure to interpersonal threat and physiological arousal through harsh, coercive, or unpredictable parent-child interactions (Chen et al., 2015; McKee et al., 2007; Whittle et al., 2022). Importantly, these behaviors do not fall along a single positive-to-negative continuum; supportive and harsh parenting practices can coexist within the same caregiving environment (Parent et al., 2016).

Emerging epigenetic evidence further suggests that caregiving experiences may contribute to biological aging. Supportive parenting has been associated with slower epigenetic aging and may buffer the biological consequences of adversity under some environmental conditions (Brody et al., 2016; Merrill et al., 2024; Metrailer et al., 2026; Sullivan et al., 2023, 2024). For example, longitudinal findings from the Future of Families and Child Wellbeing Study (FFCWB) indicated that higher observed positive parenting (e.g., warmth, responsiveness) was associated with a slower adolescent pace of aging, with the slowest aging observed among youth exposed to both higher positive parenting and lower community threat; this protective association was attenuated under conditions of high community threat (Metrailer et al., 2026). These findings suggest that caregiving may shape biological aging in context-dependent ways, but broad measures of positive parenting may obscure which specific dimensions of the caregiving environment are most consequential for biological embedding.

## The Current Study

The present study extends the Family Stress Model into the domain of biological embedding by examining whether parenting processes serve as proximal pathways through which material hardship is associated with children’s epigenetic aging. Specifically, the study tests a developmental pathway in which material hardship is associated with greater parenting stress, which in turn predicts three distinct dimensions of parenting: warmth, behavioral control, and hostility. These parenting dimensions are then examined as unique predictors of EAA and pace of aging. By distinguishing among emotional support, behavioral structure, and punitive or harsh caregiving, the present study moves beyond broad positive-versus-negative parenting composites to test whether qualitatively different caregiving behaviors are differentially associated with biological change.

Consistent with the Family Stress Model, greater material hardship was hypothesized to be associated with greater parenting stress. Greater parenting stress was expected to be associated with lower warmth, greater behavioral control and greater hostility. In turn, greater warmth and behavioral control were hypothesized to be associated with lower EAA and a slower pace of aging, whereas greater hostility was hypothesized to be associated with EAA and a faster pace of aging. Because PedBE and DunedinPACE index conceptually distinct dimensions of epigenetic aging, associations were examined separately across both outcomes rather than assuming equivalent effects across epigenetic measures. Finally, indirect effects were tested to evaluate whether material hardship was associated with epigenetic aging through parenting stress and subsequent caregiving behaviors, consistent with a developmental pathway of biological embedding.

## Methods

### Participants

Data were drawn from the FFCWB study, an ongoing longitudinal study of 4,898 families recruited from 20 large U.S. cities (Reichman et al., 2001).Families were recruited following the focal child’s birth between 1998 and 2000, with nonmarital births oversampled at a ratio of approximately 3:1. This sampling design resulted in a cohort enriched for single-parent and socioeconomically disadvantaged families. The present study included 1,971 children with epigenetic data available at age 9. Data collected when children were ages 3 and 5 were used to assess material hardship, parenting stress, and parenting behaviors, and age-9 saliva samples were used to assess epigenetic aging. Full demographic information can be found in Table 1.

**Table 1.** Sociodemographic Characteristics and Variable Descriptives.

| **Variable** |  | **n** | **%** |
|---|---|---|---|
| **Child sex** |  |  |  |
| Male |  | 992 | 50.3 |
| Female |  | 979 | 49.7 |
| **Child race/ethnicity** |  |  |  |
| White |  | 445 | 22.6 |
| Black |  | 935 | 47.4 |
| Hispanic |  | 517 | 26.2 |
| Other |  | 70 | 3.6 |
| Missing |  | 4 | 0.2 |
| **Maternal education** |  |  |  |
| Less than high school |  | 616 | 31.3 |
| High school graduate |  | 607 | 30.8 |
| Some college |  | 520 | 26.4 |
| College graduate |  | 224 | 11.4 |
| Missing |  | 4 | 0.2 |
| **Variable** | **Mean** | **SD** | **Range** |
| **Material Hardship** | 1.47 | 1.50 | 0 - 8 |
| **Parenting Stress** | 2.19 | .68 | 1 - 4 |
| **Warmth** (Year 3) | 2.57 | .81 | 0 - 3 |
| **Warmth** (Year 5) | 3.55 | 1.55 | 0 - 5 |
| **Behavioral Control** (Year 3) | 4.23 | 1.35 | 0 - 6 |
| **Behavioral Control** (Year 5) | 4.38 | 1.30 | 0 - 6 |
| **Hostility** (Year 3) | 2.28 | 1.31 | 0 - 6 |
| **Hostility** (Year 5) | 2.28 | 1.31 | 0 - 6 |

### Procedure

Data for the present study were collected at birth and when the focal children were approximately 3 (2001–2003), 5 (2003–2006), and 9 (2007–2010) years of age. At each wave, primary caregivers completed in-home assessments addressing family and household characteristics, health, caregiving experiences, and parenting practices. Trained interviewers also completed observational assessments of the parent-child environment. At age 9, saliva samples were collected from focal children for DNA methylation analysis. All original FFCWB data-collection procedures were overseen by the Princeton University Institutional Review Board.

### Measures

#### Material Hardship

Material hardship was assessed when children were 3 years old using items derived from the Basic Needs–Ability to Meet Expenses section of the Survey of Income and Program Participation, the 1996 Panel Wave 8 Adult Well-Being Topical Module Questionnaire, the 1997 and 1999 New York City Social Indicators Surveys (Social Indicators Survey Center, 1997, 1999), and measures developed for the Family Well-Being and Welfare Reform in Iowa study. Primary caregivers responded to 10 dichotomous items assessing financial and housing-related difficulties experienced during the previous year, including difficulties paying bills, food insecurity, housing instability, and unmet basic needs. Items were coded such that higher scores indicated greater hardship and were modeled as indicators of a latent material hardship construct.

Preliminary confirmatory factor analyses (CFAs) were conducted using a diagonally weighted least squares estimator. Two rare and conceptually distinct items were excluded because of weak factor loadings and poor contribution to the measurement model (see the Supplementary Appendix for more details). The final latent construct included eight indicators. In the final measurement model, the final latent material hardship factor demonstrated adequate fit (CFI = .97, TLI = .96, RMSEA = 0.04, SRMR = 0.08) with factor loading ranging from .39 to .76.

#### Parenting Stress

Parenting stress was assessed when children were 5 years old using four items derived from the Child Development Supplement of the Panel Study of Income Dynamics. Items assessed caregivers’ feelings of being overwhelmed, frustrated, or burdened by parenting responsibilities and were rated on a 4-point Likert-type scale ranging from 1 (strongly agree) to 4 (strongly disagree). Items were reverse coded as needed so that higher scores reflected greater parenting stress. The one factor model demonstrated adequate fit (CFI = .99, TLI = .98, RMSEA = 0.07, SRMR = 0.02) with factor loadings ranging from .61 to .74.

#### Parenting Dimensions

Three conceptually distinct dimensions of parenting—warmth, behavioral control, and hostility—were examined at ages 3 and 5. Age-3 parenting constructs were included as baseline controls for their corresponding age-5 dimensions, allowing age-5 parenting to reflect variation above and beyond earlier levels of the same caregiving behavior. The final SEM therefore included separate latent factors for warmth, behavioral control, hostility at both developmental waves.

#### Warmth

Warmth was assessed through interviewer observations derived from the Home Observation for Measurement of the Environment (HOME) Scale (Bradley & Caldwell, 1984). The HOME assesses characteristics of the caregiving environment through standardized observations made during the in-home assessment. Items were coded dichotomously to indicate whether specific supportive parent-child behaviors were observed. Three indicators were available at age 3 and five indicators were available at age 5. Higher scores reflected greater observed parental warmth and responsiveness. Standardized factor loadings ranged from .83 to .93 at age 3 and from .76 to .90 at age 5, supporting the coherence of a latent warmth construct. The final SEM specifically modeled the three age-3 indicators and five age-5 indicators as separate latent warmth factors.

#### Behavioral Control and Hostility

Behavioral control and hostility were assessed using items from the Parent-Child Conflict Tactics Scale (CTS-PC; Straus et al., 1998). Primary caregivers reported how frequently they had used each parenting behavior during the previous year on a 7-point scale ranging from 0 (never) to 6 (more than 20 times). Behavioral control items reflected non-harsh strategies used to structure and guide children’s behavior, whereas hostility items reflected harsh, coercive, or psychologically aggressive responses. Four indicators were used to define behavioral control, and six indicators were used to define hostility at both ages 3 and 5. Standardized factor loadings ranged from .60 to .77 for behavioral control and from .57 to .80 for hostility at age 3. At age 5, loadings ranged from .66 to .78 for behavioral control, with the hostility items similarly demonstrating substantive loadings on their intended factor.

The distinction between behavioral control and hostility was supported by a preliminary CFA. A two-factor model separating the parenting dimensions fit significantly better than a one-factor model in which all items were constrained to load on a common parenting factor, Δχ²(1) = 356.34, p < .001, supporting their treatment as related but distinct caregiving dimensions.

#### Epigenetic Aging

Saliva samples were collected during age-9 in-home visits by Westat Inc., the FFCWS data-collection subcontractor, using Oragene® DNA Self-Collection kits (OGR-500; DNA Genotek Inc.). DNA methylation was assayed using the Infinium HumanMethylation450 BeadChip and Infinium MethylationEPIC BeadChip arrays (Illumina) according to manufacturer protocols. Quality-control procedures excluded samples with outlying methylation or bisulfite-conversion values. Cell-type proportions were estimated from saliva DNA methylation data using established reference-based procedures for pediatric samples (Middleton et al., 2022). Two complementary measures of epigenetic aging were examined: PedBE epigenetic age acceleration and DunedinPACE.

#### PedBE Epigenetic Age Acceleration (EAA)

EAA was assessed using the Pediatric Buccal Epigenetic Clock (PedBE; McEwen et al., 2020), an age-based methylation estimator developed specifically for pediatric oral tissue. PedBE EAA was operationalized as the residual variation in estimated methylation age after accounting for chronological age, such that positive values indicated an older-than-expected methylation profile and negative values indicated a younger-than-expected profile. Buccal epithelial cell composition was incorporated into the estimation procedure as recommended.

#### DunedinPACE

Pace of biological aging was assessed using DunedinPACE (Belsky et al., 2022), a DNA methylation biomarker developed from longitudinal change across multiple physiological systems. Unlike age-based clocks, DunedinPACE estimates the rate of biological change rather than deviation between estimated and chronological age. Values are scaled relative to a pace of approximately 1 biological year per chronological year, with higher values reflecting a faster estimated pace of aging. The present study used the most recent DunedinPACE specification available in the FFCWB dataset, *PoAm45*.

## Statistical Analysis

Structural equation modeling (SEM) was conducted in R using the *lavaan* package to evaluate the hypothesized developmental pathway linking material hardship, parenting stress, parenting dimensions, and epigenetic aging. Models were estimated using the weighted least squares mean- and variance-adjusted (WLSMV) estimator because the latent measurement model included categorical and ordinal indicators. Pairwise present data were used to accommodate missingness. The final analytic model included 1,971 participants.

Latent variables were specified for material hardship, parenting stress, warmth, behavioral control, and hostility. Parenting behaviors were modeled at ages 3 and 5, with each age-5 parenting dimension regressed on its corresponding age-3 factor to account for earlier levels of the same behavior. Parenting stress was regressed on material hardship, and age-5 warmth, behavioral control, and hostility were regressed simultaneously on parenting stress and material hardship. PedBE EAA and DunedinPACE at age 9 were then regressed simultaneously on all three age-5 parenting dimensions, parenting stress, and material hardship. Child sex and estimated buccal epithelial cell proportion were included as covariates. Residual covariances were estimated among the age-5 parenting factors and between the two epigenetic aging outcomes.

Model fit was evaluated using the comparative fit index (CFI), Tucker-Lewis index (TLI), root mean square error of approximation (RMSEA), and standardized root mean square residual (SRMR). CFI and TLI values of .90 or greater and RMSEA and SRMR values of .08 or lower were interpreted as indicating acceptable model fit, with CFI and TLI values approaching .95 and RMSEA values approaching .05 indicating stronger fit. Indirect effects were estimated to evaluate several components of the hypothesized developmental pathway. First, indirect associations between material hardship and each age-5 parenting dimension through parenting stress were estimated. Second, serial indirect effects tested whether material hardship was associated with each epigenetic aging outcome through parenting stress and subsequent warmth, behavioral control, or hostility. Additional indirect effects tested pathways from material hardship through parenting behavior without parenting stress and pathways through parenting stress without parenting behavior. Statistical significance of indirect effects was evaluated using model-based confidence intervals.

## Results

### Preliminary Analyses and Model Fit

The hypothesized structural equation model demonstrated acceptable-to-good fit to the data, χ²(867) = 2198.54, p < .001, CFI = .94, TLI = .94, RMSEA = .03, 90% CI [.026, .029], and SRMR = .06. Although the chi-square test was significant, this is common in large samples; the remaining fit indices indicated that the proposed model adequately represented the observed data.

### Structural Associations

As hypothesized, greater material hardship was associated with greater parenting stress, β = .35, p < .001. In turn, greater parenting stress was associated with greater behavioral control, β = .10, p = .008, greater hostility, β = .20, p < .001, and lower warmth, β = −.21, p < .001, after accounting for earlier levels of each parenting dimension. Material hardship was not directly associated with any age-5 parenting dimension once parenting stress and baseline parenting were included in the model.

Parenting dimensions showed distinct associations with children’s epigenetic aging. Greater behavioral control was associated with a slower DunedinPACE, β = −.10, p < .001, and greater warmth was similarly associated with a slower pace of aging, β = −.06, p = .007. In contrast, greater hostility was associated with a faster DunedinPACE, β = .07, p = .011. Hostility was also associated with greater PedBE EAA, β = .10, p = .003, whereas behavioral control and warmth were not significantly associated with PedBE. Neither material hardship nor parenting stress showed significant direct associations with either epigenetic aging outcome after accounting for parenting behaviors or covariates. A more comprehensive breakdown of these results can be found in Table 2.

**Table 2.** Direct Effects from the Primary Path Model.

| **Outcome / Predictor** | **B** | **SE** | **β** | **95% CI for B** | **p** |
|---|---|---|---|---|---|
| **Parenting Stress** |  |  |  |  |  |
| Material Hardship | .358 | .047 | .346 | [0.265, 0.450] | < **.001** |
| Child sex | -.022 | .035 | -.018 | [-0.091, 0.046] | .521 |
| **Warmth (Y5)** |  |  |  |  |  |
| Parenting Stress | -.266 | .063 | -.208 | [-0.390, -0.143] | < **.001** |
| Material Hardship | .024 | .074 | .018 | [-0.121, 0.170] | .742 |
| Warmth (Y3) | .344 | .05 | .399 | [0.246, 0.441] | < **.001** |
| Child Sex | .089 | .056 | .056 | [-0.022, 0.199] | .115 |
| **Behavioral Control (Y5)** |  |  |  |  |  |
| Parenting Stress | .119 | .045 | .097 | [0.032, 0.206] | **.008** |
| Material Hardship | .028 | .051 | .022 | [-0.073, 0.129] | .584 |
| Behavioral Control (Y3) | .623 | .039 | .585 | [0.546, 0.700] | < **.001** |
| Child sex | -.303 | .044 | -.199 | [-0.390, -0.216] | < **.001** |
| **Hostility (Y5)** |  |  |  |  |  |
| Parenting Stress | .175 | .034 | .196 | [0.108, 0.241] | < **.001** |
| Material Hardship | .038 | .039 | .042 | [-0.038, 0.115] | .325 |
| Hostility (Y3) | .627 | .045 | .648 | [0.539, 0.715] | < **.001** |
| Child Sex | -.131 | .032 | -.119 | [-0.193, -0.069] | < **.001** |
| **DunedinPACE** |  |  |  |  |  |
| Warmth (Y5) | -.013 | .005 | -.064 | [-0.022, -0.003] | **.007** |
| Behavioral Control (Y5) | -.020 | .006 | -.096 | [-0.031, -0.009] | < **.001** |
| Hostility (Y5) | .021 | .008 | .072 | [0.005, 0.037] | **.011** |
| Parenting Stress | -.008 | .005 | -.031 | [-0.018, 0.002] | .125 |
| Material Hardship | .003 | .006 | .010 | [-0.010, 0.015] | .675 |
| BEC | .923 | .014 | .772 | [0.896, 0.951] | < **.001** |
| Child Sex | .006 | .005 | .020 | [-0.003, 0.016] | .186 |
| **PedBE EAA** |  |  |  |  |  |
| Warmth (Y5) | -.005 | .022 | -.006 | [-0.048, 0.039] | .838 |
| Behavioral Control (Y5) | -.034 | .026 | -.040 | [-0.084, 0.017] | .188 |
| Hostility (Y5) | .115 | .039 | .097 | [0.038, 0.191] | **.003** |
| Parenting Stress | .047 | .025 | .044 | [-0.002, 0.096] | .062 |
| Material Hardship | -.018 | .028 | -.017 | [-0.073, 0.036] | .510 |
| BEC | -3.365 | .073 | .691 | [-3.509, -3.222] | < **.001** |
| Child Sex | .071 | .022 | .055 | [0.028, 0.115] | **.001** |

Note. BEC = buccal epithelial cell composition; EAA = epigenetic age acceleration; Y3 = year 3; Y5 = year 5; β = standardized beta coefficient

### Indirect Effects

Material hardship was indirectly associated with all three age-5 parenting dimensions through parenting stress. Greater hardship was associated with greater behavioral control, b = .043, 95% CI [.009, .076], p = .013; greater hostility, b = .062, 95% CI [.034, .091], p < .001; and lower warmth, b = −.095, 95% CI [−.144, −.047], p < .001, through greater parenting stress.

Several serial indirect pathways from material hardship to epigenetic aging were also significant. For DunedinPACE, significant indirect effects emerged through parenting stress and subsequent behavioral control, b = −.001, p = .036; hostility, b = .001, p = .030; and warmth, b = .001, p = .033. The directions of these pathways reflected the distinct parenting processes: hardship was associated with greater parenting stress, which predicted greater behavioral control and thereby a slower pace of aging; greater hostility and thereby a faster pace of aging; and lower warmth and thereby a faster pace of aging. For PedBE EAA, only the serial indirect pathway through parenting stress and hostility was significant, b = .007, 95% CI [.001, .013], p = .016. Serial indirect effects through behavioral control and warmth were not significant for PedBE EAA. A more comprehensive breakdown of significant and non-significant indirect effects can be found in Table 3.

**Table 3.** Indirect Effects of Material Hardship on Parenting Processes and Biological Aging.

| Indirect Pathway | B | SE | β | 95% CI for B | p |
|---|---|---|---|---|---|
| **MH → Stress → Parenting** |  |  |  |  |  |
| MH → Stress → Behavioral Control | .043 | .017 | .034 | [0.009, 0.076] | **.013** |
| MH → Stress → Hostility | .062 | .015 | .068 | [0.034, 0.091] | **< .001** |
| MH → Stress → Warmth | -.095 | .025 | -.072 | [-0.144, -0.047] | **< .001** |
| **MH → Stress → Parenting → Aging** |  |  |  |  |  |
| MH → Stress → Warmth → PACE | .001 | .001 | .005 | [0.000, 0.002] | **.033** |
| MH → Stress → Behavioral Control → PACE | -.001 | .000 | -.003 | [-0.002, 0.000] | **.036** |
| MH → Stress → Hostility → PACE | .001 | .001 | .005 | [0.000, 0.002] | **.030** |
| MH → Stress → Warmth → EAA | .000 | .002 | .000 | [-0.004, 0.005] | .838 |
| MH → Stress → Behavioral Control → EAA | -.001 | .001 | -.001 | [-0.004, 0.001] | .240 |
| MH → Stress → Hostility → EAA | .007 | .003 | .007 | [0.001, 0.013] | **.016** |

Note. MH = Material Hardship, EAA = epigenetic age acceleration; PACE = DunedinPACE

**Figure 1.**
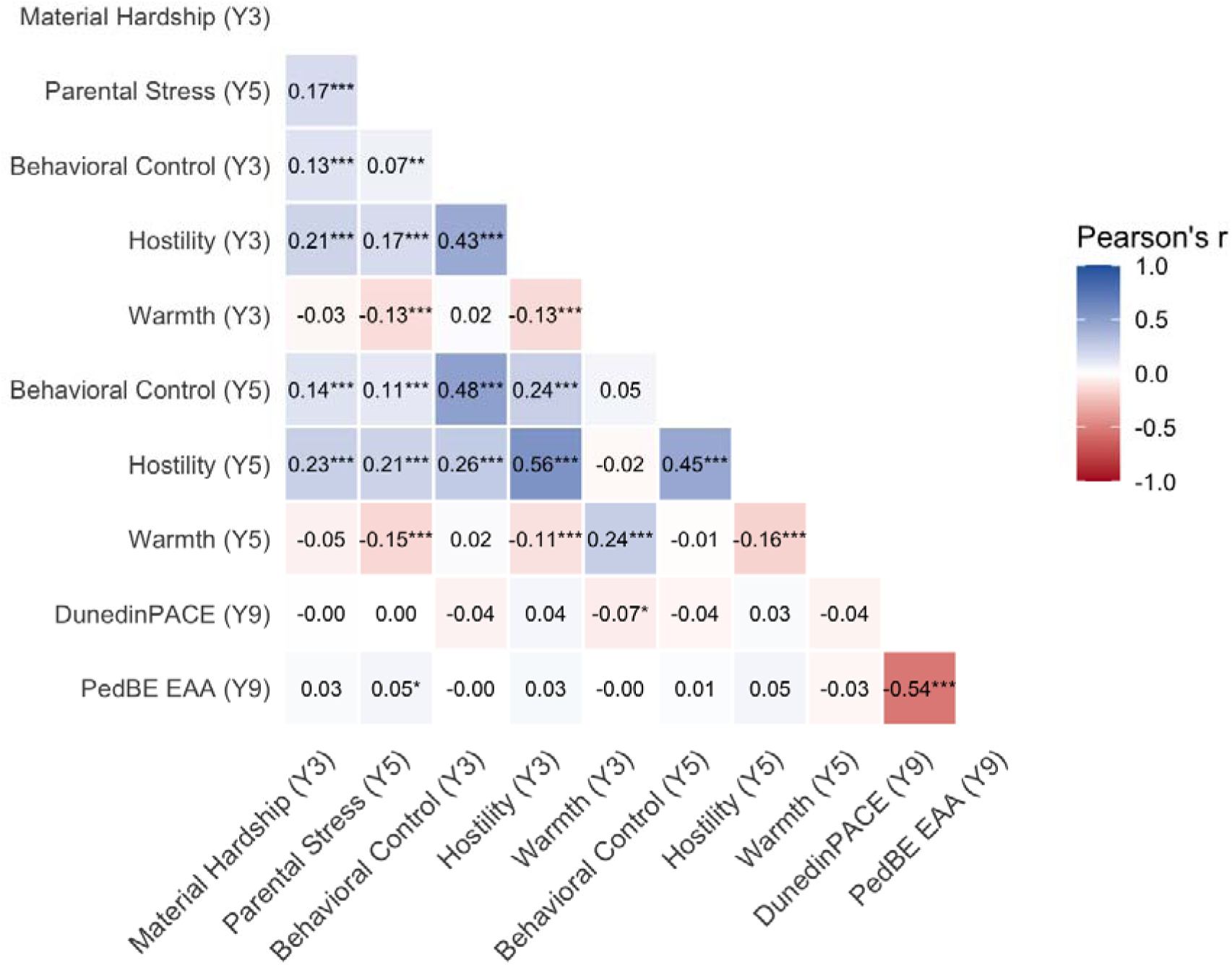
Correlation Matrix of Study Variables. Note. * p < .05, ** p < 0.01, *** p < .001

**Figure 2.**
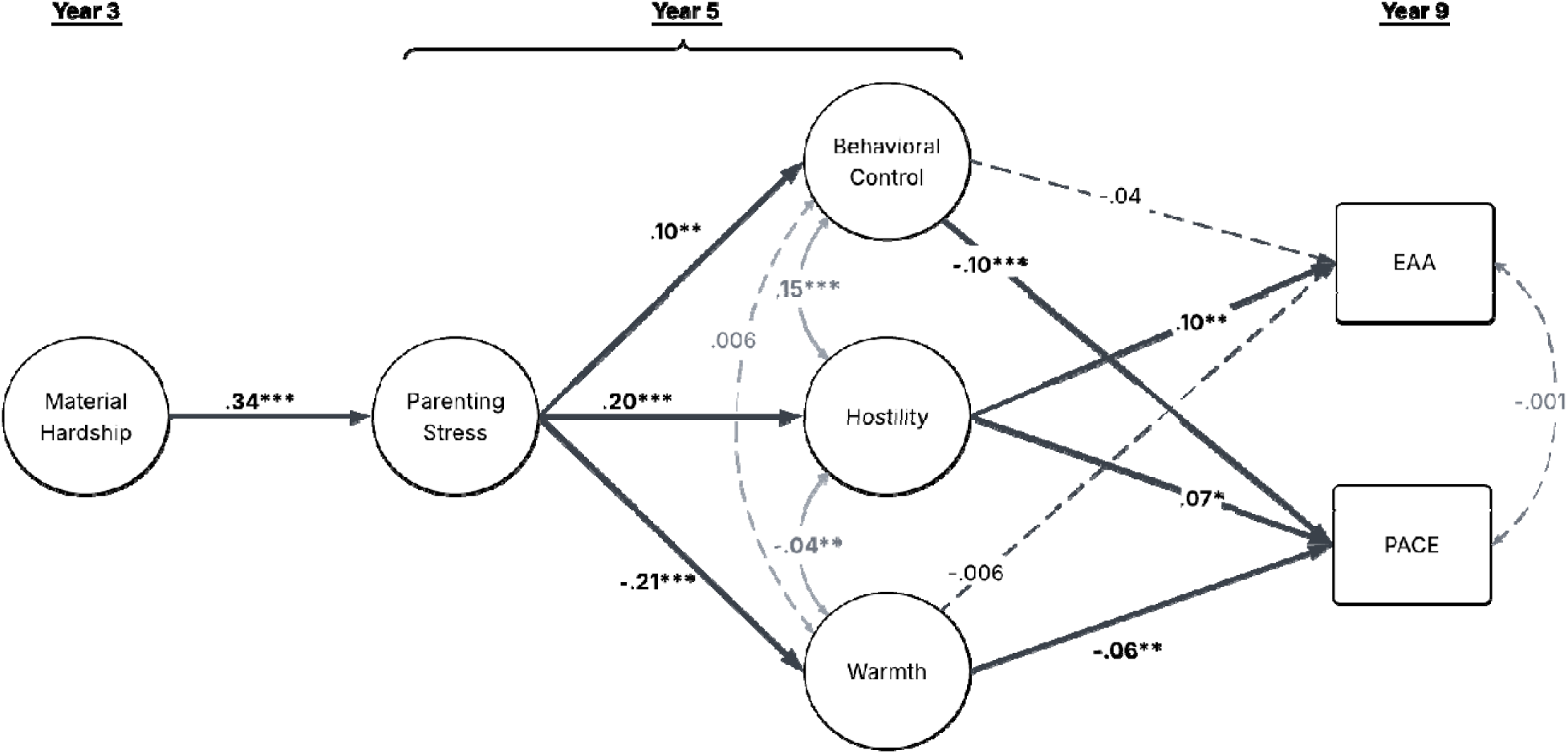
Path Model of Material Hardship on Parenting Processes and Biological Aging. Note. EAA = epigenetic age acceleration; PACE = pace of aging; * p < .05, ** p < .01, *** p < .001

## Discussion

This study examined whether family processes help explain how material hardship becomes biologically embedded across early childhood development. Guided by the Family Stress Model (Conger, 1981; Masarik & Conger, 2017), a longitudinal pathway model tested whether material hardship early in life was associated with subsequent parenting stress, whether parenting stress predicted three distinct dimensions of parenting, and whether these parenting behaviors were, in turn, associated with children’s epigenetic aging. Overall, the findings were consistent with a biologically embedded family stress model in which socioeconomic adversity was associated with children’s biological aging through proximal family processes rather than through direct associations alone. More specifically, material hardship increased parental stress and disrupted parenting practices that were associated with accelerated epigenetic aging. Together, these findings extend the Family Stress Model by positioning parental stress and parenting as proximal, modifiable pathways through which broader socioeconomic adversity may become biologically embedded during childhood.

### Material Hardship & Parenting Stress

Consistent with the Family Stress Model, greater material hardship during early childhood was associated with greater parenting stress two years later. Experiences of material hardship have been shown to place repeated demands on parents by increasing uncertainty, constraining available resources, and requiring parents to manage competing financial and caregiving demands. Accordingly, the association between material hardship and parenting stress is consistent with longstanding evidence that economic pressure affects children’s development partly through altering parental emotional, temporal, and regulatory resources (K. A. Crnic, 2024; K. Crnic & Ross, 2017; Gagnon et al., 2026; Masarik & Conger, 2017). However, this pathway should not be interpreted as situating the developmental consequences of socioeconomic adversity primarily within parents. Instead, parenting stress should be understood as a process that unfolds within a broader ecology characterized by structural and material constraints, as well as the broader contexts surrounding the family. In this sense, parenting stress represents one potential point of transmission between distal socioeconomic conditions and children’s proximal developmental environments.

### Parenting Stress, Multidimensional Parenting, & Epigenetic Aging

Parenting stress was subsequently associated with multiple dimensions of parenting behavior, including greater hostility, lower warmth, and greater behavioral control. Heightened parenting stress may reduce parents’ capacity to regulate affect and behavior during challenging parent-child interactions (Barroso et al., 2018; Deater-Deckard, 1998; Jackson & Choi, 2018). Parents under significant stress have been shown to rely more on reactive, coercive, and harsh disciplinary responses when attempting to regulate their child’s behavior, and research consistently finds these parenting behaviors to be consequential to children’s social, psychological, and biological development (Brody et al., 2014; Burt et al., 2021; Jackson & Choi, 2018).

Importantly, in the present study, hostility was subsequently associated with both a faster pace of biological aging and a greater EAA, making it the parenting dimension most consistently associated with biological aging across the three parenting dimensions measured. Repeated exposure to parental hostility may, therefore, contribute to a more chronically demanding stress-regulatory environment as children attempt to navigate moments of conflict under heightened physical arousal. Consistent with this interpretation, prior work has linked punitive and harsh discipline with alterations in children’s stress-response and neurodevelopmental systems (Chen et al., 2015; Whittle et al., 2022).

In contrast to the stress-related risk associated with hostility, warmth represented a distinct dimension of the caregiving environment that was associated with slower biological aging. Greater parenting stress was associated with lower observed warmth, suggesting that the consequences of stress for parenting behavior and subsequent biological aging may extend beyond increases in hostile or punitive behavior to reductions in supportive and emotionally responsive interactions. Warmth may be particularly relevant for children’s psychosocial and biological development because responsive parenting can provide a predictable source of safety and co-regulation during early childhood (Bornstein et al., 2026; Carroll et al., 2013; Padilla-Walker et al., 2016). Repeated experiences of warmth may therefore reduce the regulatory demands placed on children as they navigate broader environmental stress, providing an interpersonal context in which children can rely on caregivers for support and regulation (Metrailer et al., 2026). At the same time, maintaining warm and responsive interactions may become increasingly difficult as parents contend with the persistent demands associated with socioeconomic adversity (Gagnon et al., 2026; Lee et al., 2024; Masarik & Conger, 2017). Thus, the association between greater parenting stress and lower warmth highlights one way in which broader economic conditions may constrain a potentially supportive feature of children’s immediate caregiving environments.

Behavioral control (e.g., redirection, taking away privileges, utilizing time-outs) may be another parenting dimension that protects against the biological embedding of adversity, particularly for families facing socioeconomic hardship (Highlander et al., 2025). Greater parenting stress was associated with greater behavioral control which, in turn, was associated with a *slower* pace of aging. This finding further challenges a characterization of parenting under stress as uniformly compromising and highlights the importance of distinguishing behavioral structure from coercive or psychologically controlling forms of parenting (Bean et al., 2006; Bornstein et al., 2026; Bornstein & Bradley, 2012). For instance, behavioral control may function differently from harsh and punitive discipline because it involves parents’ efforts to establish expectations, monitor behavior, and maintain consistent boundaries rather than relying on coercion or hostility (Barber et al., 1994). Under conditions of heightened family stress, greater structure may provide children with predictability and behavioral guidance within an otherwise uncertain environment.

From this perspective, the positive association between parenting stress and behavioral control may reflect parents’ efforts to increase structure in response to contextual demands, rather than simply another manifestation of disrupted caregiving (Bornstein & Bradley, 2012; Putnick et al., 2008). Consistent with this possibility, research examining parenting in disadvantaged environments suggests that behavioral control, when characterized by clear expectations, guidance, and supportive limit setting, may represent an adaptive caregiving strategy that promotes children’s regulation and adjustment (Bocknek et al., 2020; Brody & Flor, 1998; Goldner et al., 2016). Evidence from Behavioral Parent Training programs further underscores the potential benefits of effective behavioral management practices. For example, Merrill et al. (2024) found that children receiving internet-delivered Parent–Child Interaction Therapy (PCIT), which emphasizes positive reinforcement and consistent behavioral management, exhibited a slower epigenetic pace of aging. Within this context, the association between behavioral control and a slower pace of aging in the present study raises the possibility that predictable behavioral structure may represent one component of the caregiving environment that supports children’s regulation under adversity beyond parental warmth and support.

### Developmental Coupling of Bioecological Processes

The pattern of indirect rather than direct associations between material hardship and epigenetic aging may also inform dimensional models of childhood adversity. The threat– deprivation framework distinguishes experiences involving the absence of expected environmental inputs from those involving threat of harm and proposes that these dimensions influence development through partially distinct mechanisms (McLaughlin et al., 2014, 2021). Prior research on biological aging has similarly suggested that threat-related experiences may be more consistently associated with accelerated aging than deprivation-related experiences (Hogan et al., 2024; Sumner et al., 2019). However, the present findings raise the possibility that distal deprivation-related conditions may alter family processes in ways that subsequently shape parenting practices that reflect both threat (e.g., hostility, punitive discipline) and deprivation-related (e.g., lower warmth) domains. Specifically, material hardship was associated with greater subsequent parenting stress, which was in turn associated with greater hostility, lower warmth, and greater behavioral control. Thus, material hardship may represent one deprivation-related starting point in a developmental sequence rather than being directly or immediately associated with child biological embedding.

Building on bioecological, transactional, and developmental systems prospectives (Bronfenbrenner, 1977; Sameroff, 2000; Smith & Thelen, 2003), we conceptualize this process as **developmental coupling of bioecological processes**, whereby theoretically distinct dimensions of adversity may become interconnected through ecological and relational processes that shape children’s exposure to both adversity and protection across development. Importantly, this perspective does not suggest that deprivation “becomes” threat or that these dimensions operate through identical developmental mechanisms. Rather, distal conditions associated with one dimension of adversity (e.g., SES-related deprivation) may alter family processes, which in turn shape the frequency and intensity of children’s exposure to proximal forms of adversity (e.g., threat-related parenting practices) or protection (e.g., behavioral control), with potentially distinct implications for biological and psychosocial development. Although parenting dimensions such as hostility, warmth, and behavioral control may arise from a shared ecological condition (e.g., material hardship), they contribute to divergent developmental pathways, particularly with respect to biological aging. In the present study, although material hardship indirectly increased hostility and behavioral control, hostility was associated with increased EAA and a faster pace of aging, whereas behavioral control was associated with a slower pace of aging, suggesting a potentially protective function. These findings raise the possibility that adversity may initiate interconnected developmental processes that simultaneously increase vulnerability while maintaining or strengthening sources of protection.

The developmental coupling of bioecological processes perspective may help bridge dimensional and person-centered approaches to understanding the long-term consequences of childhood adversity (Chow et al., 2025; McLaughlin et al., 2023; Sisitsky et al., 2023). Whereas dimensional approaches emphasize the distinct mechanisms through which different forms of adversity influence development, person-centered approaches characterize patterns of cooccurring adversity (Jobe-Sheilds et al., 2015; McLaughlin et al., 2014). Developmental coupling offers a complementary perspective by shifting attention toward the ecological and relational processes through which these patterns emerge, influence each other, and change across devel-opmental time. By considering how distal adversity may simultaneously influence proximal risk-enhancing and resilience-enhancing processes, this perspective suggests that the developmental consequences of adversity cannot necessarily be understood by examining individual exposures or pathways in isolation. Rather, understanding biological and psychosocial development may require consideration of how adverse and protective processes become organized within the same developmental context

### Limitations, Strengths, and Future Directions

Several limitations warrant consideration when interpreting the present findings. First, although material hardship was assessed before parenting stress and subsequent parenting behaviors, parenting stress and age-5 parenting dimensions were measured concurrently, limiting conclusions about temporal ordering. Moreover, despite the longitudinal design, a prospective longitudinal approach cannot establish causality, and unmeasured family characteristics or reciprocal influences between child behavior and parenting may contribute to the observed associations. Nevertheless, emerging experimental evidence supports the possibility that modifying children’s socioeconomic environments may influence epigenetic aging. For example, Raffington et al. (2026) found that children whose mothers were randomized to receive larger unconditional cash transfers exhibited a slower epigenetic pace of aging compared with those receiving smaller transfers, although the developmental processes underlying these effects remain unclear.

Similarly, intervention research suggests that modifying parent–child relationships may influence children’s epigenetic aging. Merrill et al. (2024) found that children randomized to internet-delivered behavioral parent training program exhibited a slower epigenetic pace of aging, whereas Sullivan et al. (2024) found lower EAA among children receiving Child–Parent Psychotherapy, a family intervention focused on strengthening protective and responsive caregiving relationships, relative to a matched community sample. Together with the present findings, these three studies highlight the potential value of experimental and intervention research examining whether changes in socioeconomic conditions and parenting behaviors independently or jointly influence children’s epigenetic aging. Future studies that integrate economic and parental stress management support and parenting practices interventions could clarify the causal processes through which socioeconomic adversity becomes biologically embedded.

An additional limitation concerns the measurement and interpretation of biological aging. DNA methylation was assessed only at age 9, precluding examination of earlier differences or within-person changes in epigenetic aging across early childhood development. Furthermore, extending this work beyond epigenetic aging algorithms to examine broader patterns of epigenetic regulation, including epigenome-wide associations and methylation signatures related to inflammation and hypothalamic-pituitary-adrenal axis functioning, may help clarify the molecular processes underlying the observed associations (Danese et al., 2011; Wesarg et al., 2020). Such an approach could provide a more comprehensive understanding of how repeated parenting experiences become biologically embedded and whether different parenting dimensions are associated with distinct patterns of epigenetic regulation.

Finally, although the FFCWB study sample includes substantial socioeconomic and demographic diversity, its sampling design and the availability of epigenetic data within a subset of participants may limit generalizability. The available measures also did not fully capture social and structural experiences that may shape parenting and children’s biological development, including but not limited to racial discrimination and culturally relevant protective processes such as racial and ethnic socialization (Del Toro et al., 2024; Timperio et al., 2015). Future research incorporating these experiences, alongside broader environmental conditions such as neighborhood disadvantage or community-level threat, could clarify how parenting processes operate within different ecological contexts. This is particularly relevant given evidence that community-level threat may attenuate the protective association between positive parenting and epigenetic aging (Metrailer et al., 2026), suggesting that the developmental significance of parenting may depend partly on the contexts in which families inhabit.

Despite these limitations, the present study has several methodological and conceptual strengths. The use of a large socioeconomically and demographically diverse longitudinal cohort enabled the examination of associations between material hardship in early childhood, subsequent family processes, and epigenetic aging in middle childhood—processes that might not be easily captured in smaller research samples and designs. The integration of parent-report, observational methods, and DNA methylation biomarkers reduced reliance on a single measurement method and the potential for shared method bias. Moreover, distinguishing among warmth, behavioral control, and hostility, rather than relying on broad positive and negative parenting composites, allowed for a more differentiated examination of caregiving processes that may have distinct associations with children’s biological development. Accounting for earlier levels of each parenting dimension further strengthened the longitudinal analysis by examining subsequent parenting behaviors beyond their corresponding age-3 levels. Finally, incorporating both PedBE and DunedinPACE allowed the study to examine whether hypothesized family processes were associated with epigenetic indicators developed to capture different aspects of biological aging.

## Conclusion

In conclusion, the present study extends the Family Stress Model into the domain of biological embedding by identifying distinct family processes through which material hardship is associated with children’s biological development. Greater material hardship was associated with elevated parenting stress, which in turn was associated with lower warmth, greater behavioral control, and greater hostility. These parenting dimensions demonstrated contrasting associations with children’s epigenetic aging, with warmth and behavioral control associated with a slower pace of aging and hostility associated with both a faster pace of aging and greater EAA. Importantly, the significant serial indirect pathways suggest that socioeconomic adversity may become biologically embedded through intervening family processes rather than operating solely through direct associations with children’s epigenetic outcomes. Understanding the biological embedding of adversity may, therefore, require examining not only the consequences of individual environmental and interpersonal exposures, but also the developmental processes through which these experiences become interconnected across time. Through identifying parenting, a potentially modifiable component of these pathways, the present findings provide a foundation for future research examining how economic and parenting interventions may jointly support children’s biological development.

## Data Availability

All data produced are available online at https://ffcws.princeton.edu/

## APPENDICES

**Table A1.** Description of items used in final measurement model.

| Construct | Year | Item Indicator | Question | Respondent | Coding |
|---|---|---|---|---|---|
| Material Hardship | 3 | m3i23a | In past year, did you receive free food or meals? | Caregiver | 0 = No<br>1 = Yes |
|  |  | m3i23b | In past year, did you not pay full rent or mortgage? |  |  |
|  |  | m3i23c | In past year, were you evicted from home? |  |  |
|  |  | m3i23d | In past year, did you not pay full gas/oil/electricity bill? |  |  |
|  |  | m3i23e | In past year, did you borrow money from friends/family? |  |  |
|  |  | m3i23f | In past year, did you move in with other people because of financial problems? |  |  |
|  |  | m3i23h | In past year, did someone not see doctor/hospital? |  |  |
|  |  | m3i23i | In past year, did you cut back on buying yourself clothes? |  |  |
| Parenting Stress | 5 | m4b6a | Being a parent is harder than I thought it would be | Caregiver | 1 = Strongly Agree<br>2 = Agree<br>3 = Disagree<br>4 = Strongly Disagree<br>\**Items reverse coded analysis* |
|  |  | m4b6b | I feel trapped by my responsibilities as a parent |  |  |
|  |  | m4b6c | Taking care of my child(ren) is much more work than pleasure |  |  |
|  |  | m4b6d | I often feel tired/worn out/exhausted from raising a family |  |  |
| Warmth | 3 | o3t7 | Parent spontaneously praises child at least twice | Interviewer | 0 = Did not occur<br>1 = Did occur |
|  |  | o3t8 | Parent’s voice conveys positive feelings toward child |  |  |
|  |  | o3t9 | Parent caressed or kissed child at least once |  |  |
|  | 5 | o4t4 | Parent helped child demonstrate some achievement or mentioned strength |  |  |
|  |  | o4t5 | Parent spontaneously praised child's behavior or qualities twice |  |  |
|  |  | o4t6 | Parent used some term of endearment twice or more during visit |  |  |
|  |  | o4t7 | Parent's voice conveyed positive feelings when speaking of or to child |  |  |
|  |  | o4t8 | Parent caressed or kissed child at least once |  |  |
| Behavioral Control | 3 | p3j1 | Past year, times PCG explained to child why something child did was wrong | Caregiver | 0 = This never happened<br>1= Once<br>2 = Twice<br>3 = 3-5 times<br>4 = 6-10 times<br>5 = 11-20 times<br>7 = Yes, but not in the past year (recoded as 0) |
|  |  | p3j2 | Past year, times PCG put child in time out or sent child to room |  |  |
|  |  | p3j5 | Past year, times PCG gave child something else to do instead |  |  |
|  |  | p3j12 | Past year, times PCG took away privileges from child |  |  |
|  | 5 | p4g1 | Past year, times PCG explained to child why something child did was wrong |  |  |
|  |  | p4g2 | Past year, times PCG put child in time out or sent child to room |  |  |
|  |  | p4g5 | Past year, times PCG gave child something else to do instead |  |  |
|  |  | p4g12 | Past year, times PCG took away privileges from child |  |  |
| Hostility | 3 | p3j4 | Past year, times PCG hit child on the bottom with belt or hard object | Caregiver | 0 = This never happened<br>1= Once<br>2 = Twice<br>3 = 3-5 times<br>4 = 6-10 times<br>5 = 11-20 times<br>7 = Yes, but not in the past year (recoded as 0) |
|  |  | p3j6 | Past year, times PCG shouted, yelled, or screamed at child |  |  |
|  |  | p3j7 | Past year, times PCG spanked child on bottom with bare hand |  |  |
|  |  | p3j8 | Past year, times PCG swore or cursed at child |  |  |
|  |  | p3j10 | Past year, times PCG threatened to spank/hit child but did not |  |  |
|  |  | p3j11 | Past year, times PCG slapped child on the hand, arm, or leg |  |  |
|  | 5 | p4g4 | Past year, times PCG hit child on the bottom with belt or hard object |  |  |
|  |  | p4g6 | Past year, times PCG shouted, yelled, or screamed at child |  |  |
|  |  | p4g 7 | Past year, times PCG spanked child on bottom with bare hand |  |  |
|  |  | p4g 8 | Past year, times PCG swore or cursed at child |  |  |
|  |  | p4g10 | Past year, times PCG threatened to spank/hit child but did not |  |  |
|  |  | p4g11 | Past year, times PCG slapped child on the hand, arm, or leg |  |  |

**Table A2.** CFA fit statistics for initial and final measurement specifications.

| **Measurement Model** | **χ<sup>2</sup>** | **df** | **CFI** | **TLI** | **RMSEA** | **SRMR** |
|---|---|---|---|---|---|---|
| Material Hardship (Initial) | 166.68 | 35 | .941 | .924 | .045 | .111 |
| Material Hardship (Final) | 80.21 | 20 | .97 | .958 | .04 | .083 |
| Parenting Stress (Final) | 17.452 | 2 | .992 | .975 | .065 | .023 |
| Warmth (Final) | 8.160 | 5 | .999 | .998 | .023 | .021 |
| Conflict Tactics (Initial) | 1558.76 | 77 | .841 | .812 | .112 | .134 |
| Conflict Tactics (Final, two factor model) | 355.93 | 34 | .962 | .95 | .079 | .058 |

**Table A3.** Factor loadings for the final CFA.

| Construct | Item Indicator | B | SE | β | P |
|---|---|---|---|---|---|
| Material Hardship | m3i23a | 1.000 | -- | .601 | -- |
|  | m3i23b | 1.268 | .106 | .763 | < .001 |
|  | m3i23c | 1.232 | .137 | .741 | < .001 |
|  | m3i23d | 1.137 | .101 | .684 | < .001 |
|  | m3i23e | 1.154 | .094 | .694 | < .001 |
|  | m3i23f | 1.014 | .097 | .610 | < .001 |
|  | m3i23h | .652 | .097 | .392 | < .001 |
|  | m3i23i | 1.103 | .099 | .663 | < .001 |
| Parenting Stress | m4b6a | 1.000 | -- | .622 | -- |
|  | m4b6b | 1.068 | .061 | .664 | < .001 |
|  | m4b6c | 1.050 | .062 | .653 | < .001 |
|  | m4b6d | 1.145 | .065 | .712 | < .001 |
| Warmth (Year 3) | o3t7 | 1.000 | -- | .927 | -- |
|  | o3t8 | .892 | .070 | .826 | < .001 |
|  | o3t9 | .893 | .071 | .828 | < .001 |
| Warmth (Year 5) | o4t4 | 1.000 | -- | .797 | -- |
|  | o4t5 | 1.125 | .050 | .896 | < .001 |
|  | o4t6 | .966 | .047 | .770 | < .001 |
|  | o4t7 | 1.098 | .057 | .875 | < .001 |
|  | o4t8 | 0.958 | .047 | .764 | < .001 |
| Behavioral Control (Year 3) | p3j1 | 1.000 | -- | .716 | -- |
|  | p3j2 | 1.077 | .048 | .772 | < .001 |
|  | p3j5 | .830 | .043 | .595 | < .001 |
|  | p3j12 | .965 | .046 | .691 | < .001 |
| Behavioral Control (Year 5) | p4g1 | 1.000 | -- | .753 | -- |
|  | p4g2 | 1.013 | .037 | .763 | < .001 |
|  | p4g5 | .871 | .035 | .658 | < .001 |
|  | p4g12 | 1.029 | .037 | .775 | < .001 |
| Hostility (Year 3) | p3j4 | 1.000 | -- | .571 | -- |
|  | p3j6 | 1.274 | .066 | .728 | < .001 |
|  | p3j7 | 1.401 | .069 | .800 | < .001 |
|  | p3j8 | .996 | .066 | .569 | < .001 |
|  | p3j10 | 1.335 | .068 | .763 | < .001 |
|  | p3j11 | 1.334 | .066 | .762 | < .001 |
| Hostility (Year 5) | p4g4 | 1.000 | -- | .552 | -- |
|  | p4g6 | 1.331 | .068 | .733 | < .001 |
|  | p4g 7 | 1.303 | .067 | .718 | < .001 |
|  | p4g 8 | 1.031 | .065 | .569 | < .001 |
|  | p4g10 | 1.315 | .067 | .725 | < .001 |
|  | p4g11 | 1.295 | .063 | .664 | < .001 |

**Table A4.**
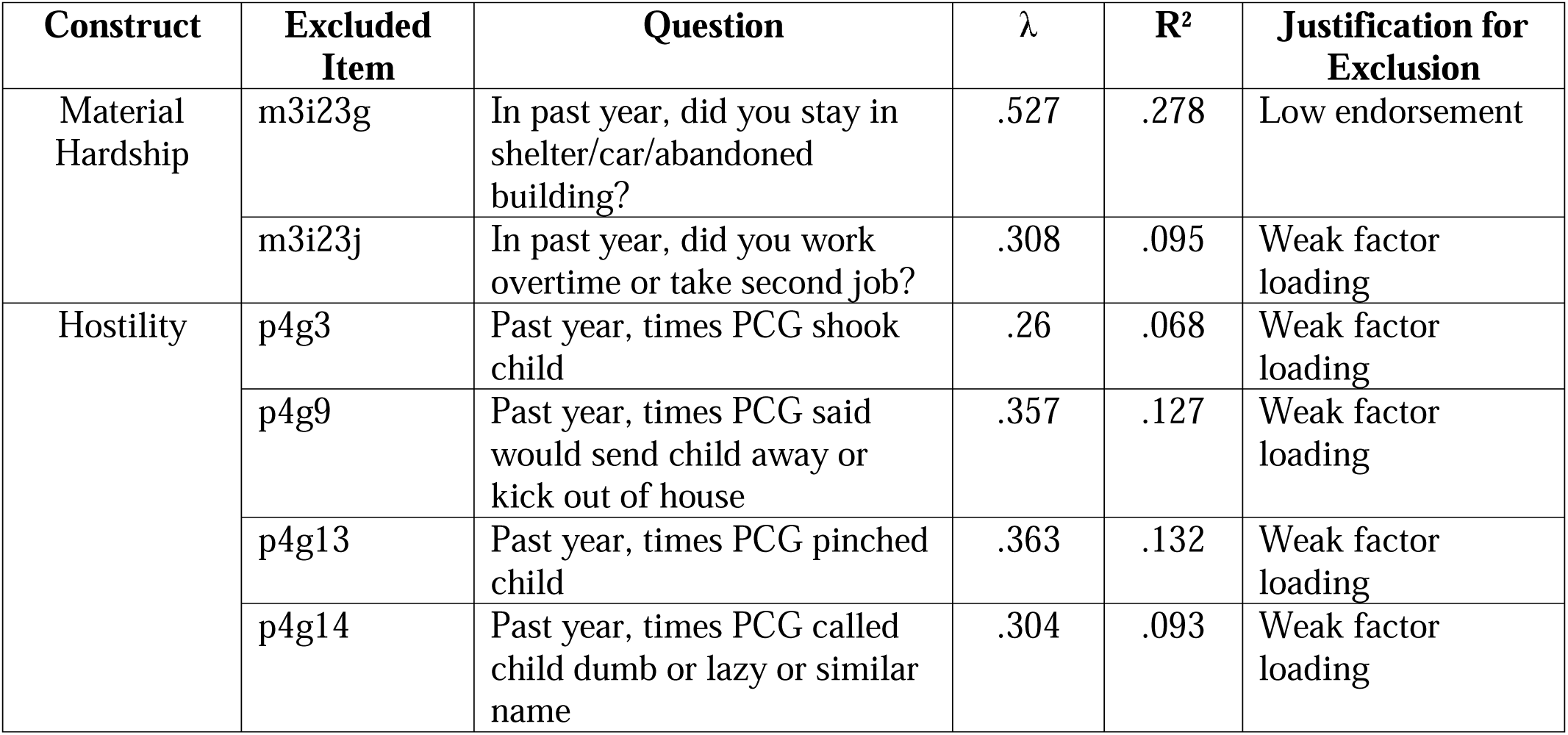
Excluded indicators and empirical or theoretical rationale for their removal.

